# Longitudinal Trajectories of Symptom Change During Antidepressant Treatment Among Managed Care Patients with Co-Occurring Depression and Anxiety

**DOI:** 10.1101/2023.09.25.23295985

**Authors:** Judith Cukor, Zhenxing Xu, Veer Vekaria, Fei Wang, Mark Olfson, Samprit Banerjee, Gregory Simon, George Alexopoulos, Jyotishman Pathak

## Abstract

Depression and anxiety are highly correlated, yet little is known about the course of each condition when presenting concurrently. This study aimed to identify longitudinal patterns and changes in depression and anxiety symptoms during antidepressant treatment, and evaluate clinical factors associated with each response pattern. Self-report Patient Health Questionnaire-9 (PHQ-9) and General Anxiety Disorder-7 (GAD-7) scores were used to track the courses of depression and anxiety respectively over a three-month window, and group-based trajectory modeling was used to derive subgroups of patients who have similar response patterns. Multinomial regression was used to associate various clinical variables with trajectory subgroup membership. Of the 577 included adults, 373 (64.6%) were women, and the mean age was 39.3 (SD: 12.9) years. Six depression and six anxiety trajectory subgroups were computationally derived; three depression subgroups demonstrated symptom improvement, and three exhibited nonresponse. Similar patterns were observed in the six anxiety subgroups. Factors associated with treatment nonresponse included higher pretreatment depression and anxiety severity and poorer sleep quality, while better overall health and younger age were associated with higher rates of remission. Synchronous and asynchronous paths to improvement were also observed between depression and anxiety. High baseline depression or anxiety severity alone may be an insufficient predictor of treatment nonresponse. These findings have the potential to motivate clinical strategies aimed at treating depression and anxiety simultaneously.

## INTRODUCTION

Symptoms of depression and anxiety often coexist. Anxious depression accounts for 42-78% of all depression cases and has been associated with higher immune and HPA-axis dysregulations and corticolimbic connectivity and more prominent cortical thinning than non-anxious depression [1]. Overall, anxious depression has poorer clinical outcomes than non-anxious depression [2]. Treatment with antidepressants is common, yet response to antidepressant treatment remains inconsistent, and dosage or medication changes often occur on a trial and error basis [3, 4].

Previous research has begun to explore trajectories of response to depression or anxiety treatment over time [5–10]. However, studies of comorbid symptom response patterns are largely limited to psychotherapy treatments [5, 7, 9], and despite the high correlation and increased functional impairment in patients with comorbid anxiety and depression symptoms, little remains known about the course of each condition when presenting together in the context of antidepressant treatment. Furthermore, many studies do not take into account the intermediate oscillations in patients’ symptoms throughout the course of the treatment period, thereby misrepresenting differences in trajectories that may exist.

One study by Smagula et al. [8] analyzed data on antidepressant response in a population of older adults from a 12 week pharmacological treatment study identified 6 distinct trajectories, including 3 responding and 3 nonresponding. Responding patients had rapid or delayed responses. A second study by Sunderland et al. [9] investigated trajectories of change for distinct depressed and anxious patients receiving internet-based cognitive behavioral therapy identified two groups with higher and lower response rates across sessions. Another study by Saunders et al. [7] investigated changes in symptoms of depression and anxiety over the course of psychotherapy identified 4 distinct trajectories of treatment response for depressive symptoms, and 5 trajectories for anxiety symptoms, including some with a delayed response. A large overlap was seen in terms of depressive and anxiety improvement, especially among those who did not respond to treatment for either diagnosis.

The most commonly identified predictor of poorer treatment response was higher baseline levels of symptom severity [6–9]. Other predictors included psychological distress [9], older age, poorer self-reported physical health and unemployment [6], longer duration of symptoms, better reported sleep, more guilt, more functional impairment [8], lower socioeconomic status, and poorer overall functioning [7].

Providing feedback to clinicians about their patients’ anticipated symptom change using expected response trajectories can improve outcomes [7], and medication decisions are informed by anticipated responses. However, the usefulness of expected trajectories may be limited if anticipated outcomes are modeled upon an averaged response curve for patients. This may reduce the precision of clinical predictions and diminish the quality of management decisions.

Understanding clinical response trajectories and their contributing factors can be instrumental in beginning to predict clinical response. It may further serve as a significant step toward personalized medicine. Compared to traditional metrics for depression which neither consider variability across multiple time points nor account for co-occurring anxiety, understanding the course of response for depression and anxiety and how they relate to each other can help further elucidate the relationship with each other [11].

Group-based trajectory modeling (GBTM), a latent class analysis method, provides a novel application for evaluating treatment response variability without depending on a prespecified remission threshold, in contrast to some prevailing literature [8, 11]. GBTM is a data-driven method that allows identification of groups of patients that share common characteristics such as demographics, comorbidities, and outcomes including treatment response [12]. Previous studies on depression have demonstrated that GBTM has the potential to capture heterogeneity during the course of illness and recovery [13, 14].

The aims of study were to identify (1) patterns (trajectories) of symptom change and (2) patient and clinical characteristics associated with increased likelihood of following the identified trajectories. To achieve this, we applied GBTM to capture the typical trajectories of symptom change throughout the initial course of antidepressant treatment in patients with co-occurring depression and anxiety. A heatmap was generated to analyze the synchronous and asynchronous relationships between the trajectories of depression and anxiety to delineate what clinicians are likely to observe in practice. In addition, the associations of baseline depression and anxiety symptom severity, sleep quality, characteristics, medical comorbidity, and overall health status with trajectory group membership were examined to identify potential predictors of symptom improvement.

## METHODS

### Overview

This cohort study used electronic health records (EHRs) and repeated Patient Health Questionnaire (PHQ-9) and Generalized Anxiety Disorder (GAD-7) measurements in 577 patients who received outpatient mental healthcare services and treatment at an urban academic medical center. We used GBTM to identify patients with similar trajectories of PHQ-9 and GAD-7 change following initiation of antidepressant therapy. These trajectory groups were then characterized using comprehensive patient information from their EHRs including demographics, comorbidities, and a variety of scoring instruments and questionnaires. The synchronicity of PHQ-9 and GAD-7 trajectories for individual patients was then described.

Finally, a multinomial regression model was built to investigate the associations of prognostic factors with trajectory group membership. The overall study design is illustrated in Figure 1.

**Figure 1.**
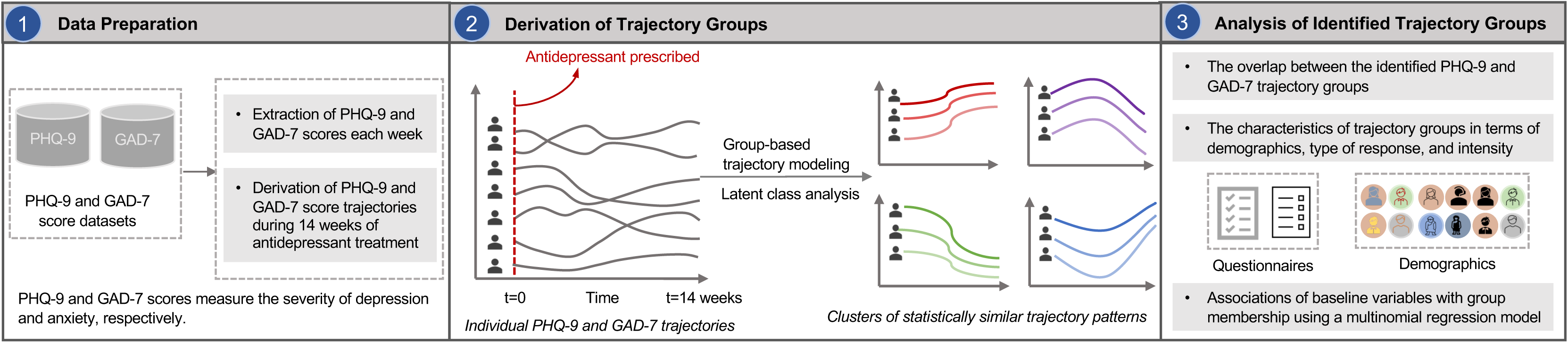
The framework of the study. Step 1: PHQ-9 and GAD-7 scores self-reported by patients each week were extracted from two datasets. Step 2: The PHQ-9 and GAD-7 scores trajectory during 14 weeks were constructed. Then, a Group-Based Trajectory Modeling (GBTM) was performed on the top of individual PHQ-9 and GAD-7 score trajectories to identify trajectory groups. Step 3: The characteristics of trajectory groups were analyzed. A multinomial regression model was built to investigate the association of baseline variables with the trajectory groups.

### Cohort Description

Individual-level EHR data were obtained from the outpatient behavioral health clinic at a major medical center in New York City. Patients at this clinic were administered multiple mental health and overall physical health related questionnaires as part of routine clinical encounters. Patients (n=577) with multiple self-reported PHQ-9 (n=3,730) and GAD-7 (n=3,704) scores between June 13, 2016 and September 26, 2020 were included. Adult patients (≥18 years old) were included in this study if a) they received antidepressant prescriptions 3 months before or up to one week after establishing care at the clinic, and b) both PHQ-9 and GAD-7 scores were measured at baseline. Prescribed antidepressants included selective serotonin reuptake inhibitors (SSRIs) (63.95%), atypical antidepressants (22.18%), serotonin and norepinephrine reuptake inhibitors (SNRIs) (9.53%), tricyclic antidepressants (TCAs) (3.99%), and monoamine oxidase inhibitors (MAOIs) (0.35%).

To ensure sufficient detail to model trajectories of depression and anxiety symptom change, patients (n=1,069) with fewer than three treatment sessions were excluded. For other patients, missing values regarding PHQ-9 and GAD-7 scores during a three-month period were imputed using the last observation carried forward (LOCF) or first observation carried backward (FOCB) method. Since a PHQ-9 score of ≥10 and GAD-7 score of ≥8 represents a reasonable cut-off for identifying probable cases of depressive disorder [7] and generalized anxiety disorder [15], respectively, patients were also excluded (n=726) if their baseline assessments were PHQ-9 <10 or GAD-7 <8. The inclusion/exclusion cascade for identifying study participants is shown in eFigure 1. IRB exemption was granted by the Weill Cornell Medicine (WCM) Institutional Review Board (IRB).

### Measures

#### Outcomes

Depression symptom severity was measured with the PHQ-9 using symptom severity ranges of absent (0-4), mild (5-9), moderate (10-14), moderately severe (15-19), or severe (20-27) [13]. Anxiety symptom severity was measured with the GAD-7 using symptom severity ranges of minimal (0-4), mild (5-9), moderate (10-14), or severe (15-21) [14]. Response was defined as a follow-up PHQ-9 (or GAD-7) score that was at least 50% lower than the patient’s baseline score [6]. Remission was defined as achieving a follow-up score of ≤5 [6].

#### Baseline characteristics

Baseline characteristics include demographic (age, gender, and race), comorbidities (post-traumatic stress disorder (PTSD), bipolar disorder, and “suicidal behavior”), and multiple validated scoring instruments and questionnaires (Alcohol Use Disorders Identification Test (AUDIT) [16], Columbia-Suicide Severity Rating Scale (C-SSRS) [17]; Pittsburgh Sleep Quality Index (PSQI) [18]; Patient-Reported Outcomes Measurement Information System (PROMIS) [19], and Sheehan Disability Scale (SDS) [20]. More details about these baseline characteristics are included in the supplementary materials.

### Statistical Analysis

Analysis was conducted in R (version 4.0.3) and GBTM [21] was performed using the package lcmm [22]. The number of groups was determined by using the Akaike Information Criterion (AIC) [23] and Bayesian Information Criterion (BIC) [24]. Lower AIC and BIC values for one model compared to another indicate better model fit. In addition, following convention with GBTM models, each class needed to contain at least 5% of the sample for it to be considered meaningful and numerically stable [7][8]. The univariate and multivariate tests were performed to obtain associations between baseline prognostic factors and trajectory groups. For the univariate analyses, we applied a student’s t-test, Mann-Whitney U test, Chi-square test, and Fisher’s exact test where appropriate. For the multivariate analyses, multinomial logistic regression models were used.

## RESULTS

### Characteristics of the derived trajectory groups

Six trajectory groups representing depressive symptom change were derived and named according to their relative baseline score and response pattern. Additional details about the process for choosing the optimal number of trajectory groups in GBTM using AIC and BIC are shown in the supplementary file (eTables 3 and 4). These included “high, nonresponse” (Cluster 1), “high, response” (Cluster 2), “moderately severe, nonresponse” (Cluster 3), “moderately severe, response” (Cluster 4), “moderate, remission” (Cluster 5), and “moderate, nonresponse” (Cluster 6). With the baseline PHQ-9 score serving as a reference point, these trajectory groups can be roughly partitioned into “severe,” “moderately severe,” and “moderate” strata, consisting of n=111 (19.24%), n=230 (39.86%), and n=236 (40.90%) patients, respectively. Within each stratum, one response and one nonresponse trajectory were observed. Within the “severe” stratum (baseline PHQ-9 ≥20), patients in the high, response group demonstrated a roughly linearly decreasing curve indicating substantial and sustained improvement in depression symptom severity overall, while high, nonresponse patients maintained a roughly constant PHQ- 9 score trajectory, indicating neither improving nor worsening symptomatology. When observing the moderately severe group (baseline PHQ-9=15-19), patients in the moderately severe, response group exhibited an inverse sigmoid-shaped curve that was initially flat, then decreased sharply and gradually tapered off. Patients in the moderately severe, nonresponse group, however, showed little to no improvement. Finally, in the moderate groups (baseline PHQ-9 <15), patients in the moderate, remission group showed an exponentially decreasing curve that reached and sustained a level below typical remission criteria (PHQ-9 ≤ 5) by the latter half of the observation window. In contrast, patients in the moderate, nonresponse group demonstrated only minimal improvement. Patient characteristics for the PHQ-9 trajectory groups are described in Table 1 and showed in Figure 2. The similar six trajectory groups on anxiety symptom change were also found, more details were included in supplementary materials.

**Figure 2.**
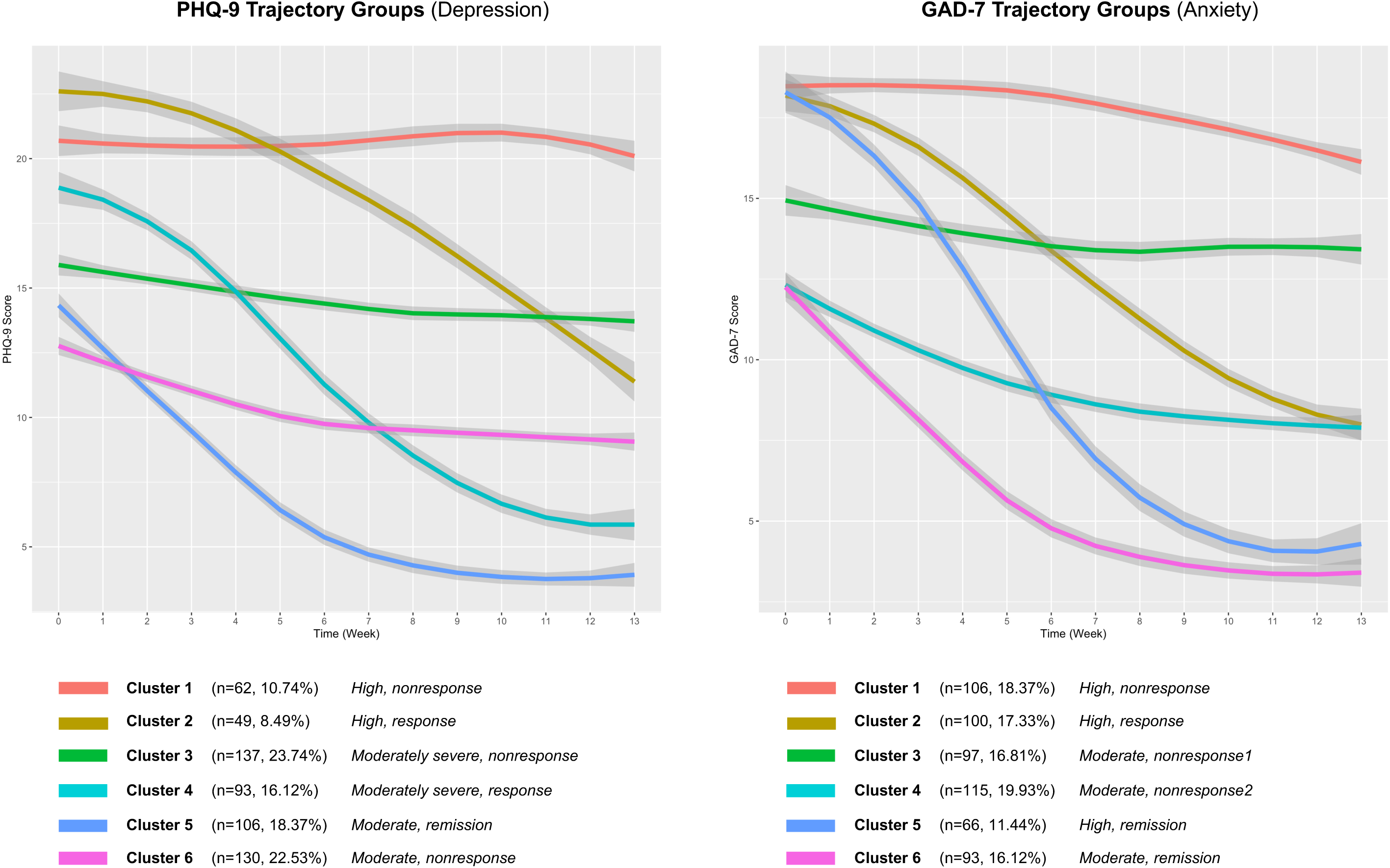
The trajectory groups derived from PHQ-9 and GAD-7 scores.

**Table 1.**
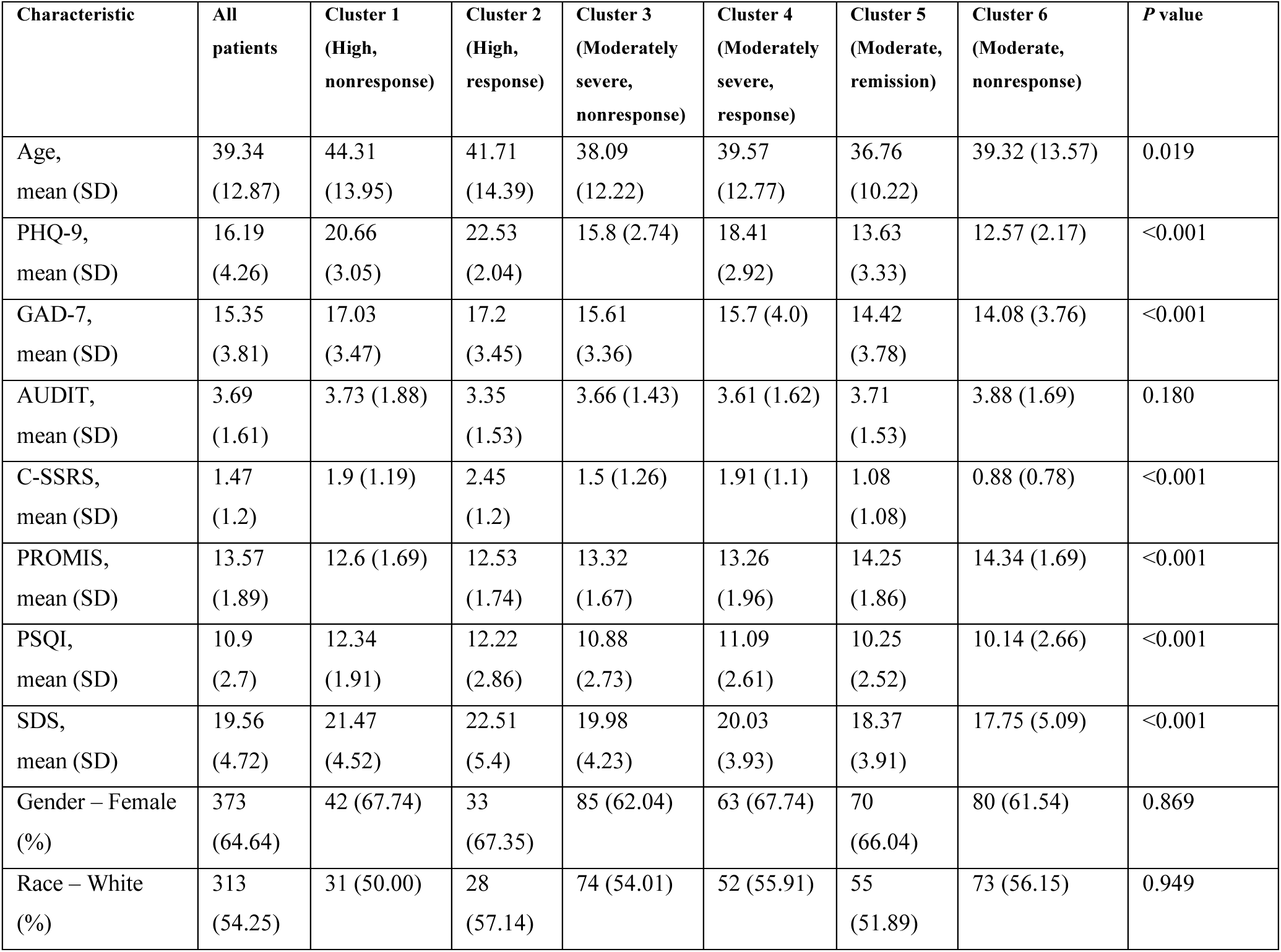
Patient features and characteristics for the PHQ-9 trajectory groups.

The overlap of patients between all permutations of the PHQ-9 and GAD-7 trajectory groups was also explored (Figure 3). The pairing with the largest degree of overlap (n=49) occurred between the PHQ-9 moderate, remission group (Cluster 5) and GAD-7 moderate, remission group (Cluster 6), suggesting similar rates of improvement for both depression and anxiety symptoms. The next largest degree of overlap (n=41) occurred between the PHQ-9 moderate, nonresponse group (Cluster 6) and GAD-7 moderate, nonresponse-2 group (Cluster 4), which suggests that a group of patients experience minimal change in depressive and anxiety symptoms during this time period. Analysis of all the overlap permutations revealed that most overlap sets (72.45% patients) displayed similar trendlines in terms of PHQ-9 and GAD-7 score change, which suggests that patients with comorbid depression and anxiety generally experience synchronous trajectories treatment response or nonresponse. However, patients in some overlap sets (27.55% patients) did demonstrate asynchronous symptom change. For example, the overlap set that contains the PHQ-9 moderately severe, nonresponse group (Cluster 3) and GAD-7 high, response group (Cluster 2) exhibited improvement in anxiety symptoms with antidepressant treatment, but similar significant improvements were not observed in depressive symptoms.

**Figure 3.**
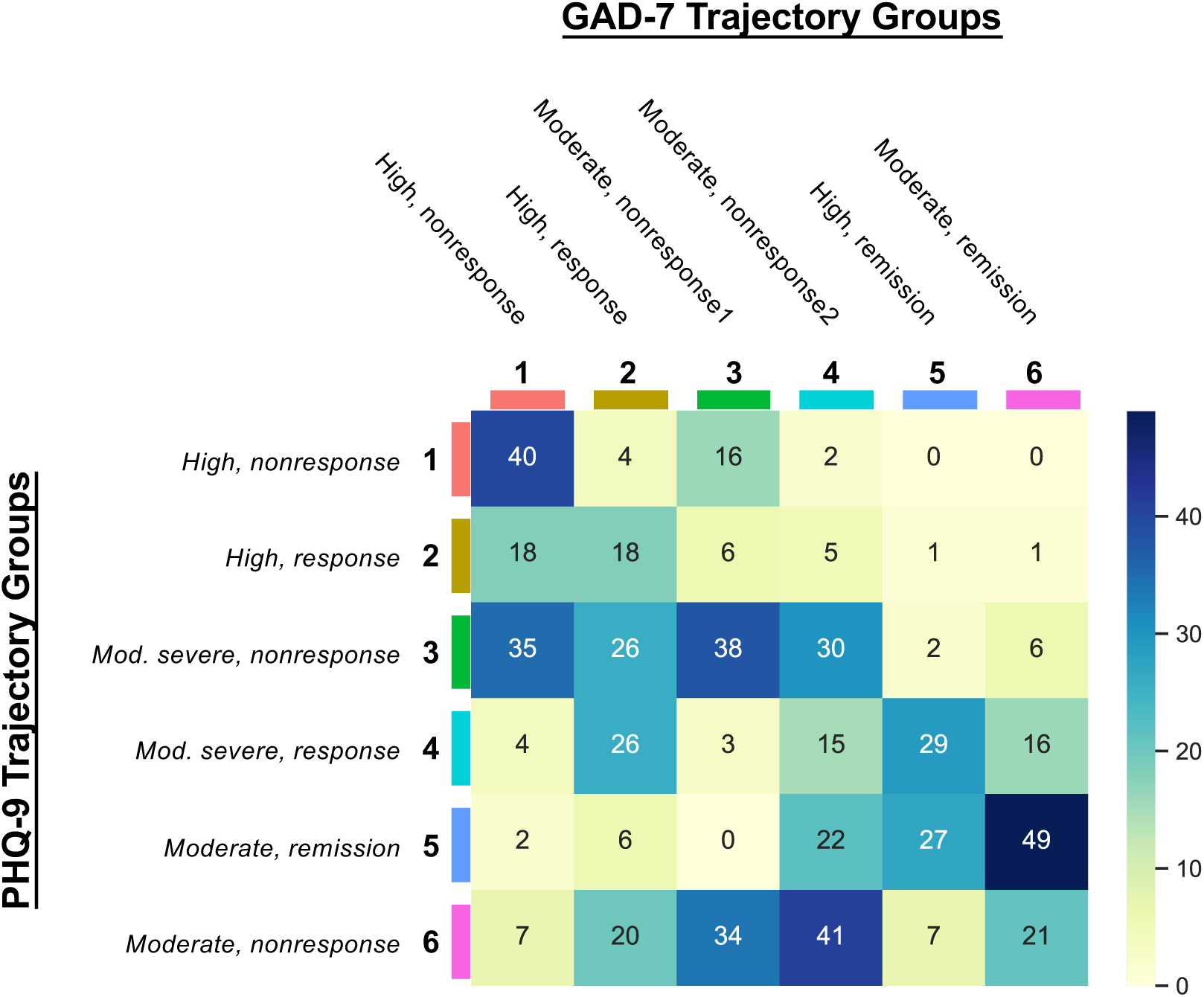
The overlap of patients between the PHQ-9 and GAD-7 trajectory groups.

### Associations of baseline patient characteristics with trajectory group membership

Associations of patients’ baseline characteristics with their derived PHQ-9 and GAD-7 trajectory classes were investigated using a multinomial regression model (see Table 2 (PHQ-9) and eTable 2 (GAD-7)) The following are probabilities in relation to being in PHQ-9 trajectory Cluster 5 (Moderate, remission): the probability of being in PHQ-9 trajectory Cluster 1 (High, nonresponse) and Cluster 2 (High, response) were higher and was associated with poorer overall health and more severity of depression prior to treatment (i.e., lower PROMIS scores and higher baseline PHQ-9 scores). The probability of being in PHQ-9 trajectory Cluster 3 (Moderately severe, nonresponse) and Cluster 4 (Moderately severe, response) relative to Cluster 5 (Moderate, remission) were higher and were associated with poorer overall health and more severity of depression prior to treatment (lower baseline PROMIS scores and higher baseline PHQ-9 scores). The probability of being in PHQ-9 trajectory Cluster 6 (Moderate, nonresponse) relative to Cluster 5 (Moderate, remission) was higher and was associated with less severity of depression prior to treatment (lower baseline PHQ-9 scores). The associations of baseline patient characteristics with GAD-7 trajectory group membership were included in the supplementary materials.

**Table 2.** Associations of baseline patient characteristics with PHQ-9 trajectory group membership.

| Characteristic | PHQ-9 Class 1<br>(High,<br>nonresponse):<br>RR (95%CI) &<br>p-value | PHQ-9 Class 2<br>(High,<br>response):<br>RR (95%CI) &<br>p-value | PHQ-9 Class 3<br>(Moderately<br>severe,<br>nonresponse):<br>RR (95%CI) &<br>p-value | PHQ-9 Class 4<br>(Moderately,<br>response):<br>RR (95%CI) &<br>p-value | PHQ-9 Class 5<br>(Moderate,<br>remission):<br>RR (95%CI) &<br>p-value | PHQ-9 Class 6<br>(Moderate,<br>nonresponse):<br>RR (95%CI)<br>& p-value |
| --- | --- | --- | --- | --- | --- | --- |
| Age | 1.0 (0.97-1.03),<br>p = .9 | 0.98 (0.95-1.02), p = .319 | 0.99 (0.97-1.02), p = .531 | 1.0 (0.97-1.02),<br>p = .819 | Reference | 1.02 (1.0-1.05),<br>p = .041 |
| Gender –<br>Female | 0.61 (0.27-1.38),<br>p = .234 | 0.58 (0.23-1.45), p = .243 | 0.7 (0.39-1.25),<br>p = .23 | 0.73 (0.37-1.44),<br>p = .361 | Reference | 0.97 (0.55-1.7),<br>p = .916 |
| Race –<br>Non-White | 1.0 (0.46-2.17),<br>p = .991 | 0.59 (0.24-1.43), p = .242 | 0.84 (0.48-1.47), p = .542 | 0.8 (0.42-1.52),<br>p = .491 | Reference | 0.94 (0.55-1.61), p = .832 |
| AUDIT | 1.04 (0.82-1.32),<br>p = .721 | 0.87 (0.65-1.18), p = .378 | 0.98 (0.81-1.18), p = .794 | 0.96 (0.78-1.19),<br>p = .723 | Reference | 1.06 (0.89-1.26), p = .519 |
| C-SSRS | 0.81 (0.54-1.22),<br>p = .311 | 1.08 (0.71-1.63), p = .716 | 0.95 (0.69-1.3),<br>p = .736 | 1.03 (0.73-1.45),<br>p = .868 | Reference | 0.87 (0.6-1.25),<br>p = .456 |
| LEC-5 | 1.08 (0.97-1.2),<br>p = .181 | 1.07 (0.93-1.22), p = .355 | 1.08 (1.0-1.16),<br>p = .05 | 1.05 (0.96-1.16),<br>p = .27 | Reference | 1.06 (0.98-1.14), p = .132 |
| PCL-C | 1.02 (0.91-1.14),<br>p = .768 | 1.02 (0.9-1.15),<br>p = .783 | 1.07 (0.98-1.16), p = .116 | 1.0 (0.91-1.1), p<br>= .967 | Reference | 0.95 (0.88-1.04), p = .258 |
| PROMIS | 0.47 (0.4-0.56),<br>p < .001 | 0.43 (0.36-0.53), p < .001 | 0.76 (0.68-0.84), p < .001 | 0.66 (0.58-0.75),<br>p < .001 | Reference | 1.07 (0.96-1.18), p = .211 |
| PSQI | 0.95 (0.79-1.12),<br>p = .525 | 0.86 (0.71-1.05), p = .147 | 0.94 (0.84-1.06), p = .316 | 0.91 (0.79-1.05),<br>p = .204 | Reference | 1.0 (0.9-1.12), p<br>= .983 |
| SDS | 0.88 (0.8-0.97),<br>p = .01 | 0.88 (0.79-0.98), p = .022 | 1.01 (0.95-1.09), p = .694 | 0.94 (0.87-1.02),<br>p = .121 | Reference | 1.0 (0.94-1.07),<br>p = .904 |
| PHQ-9 | 2.08 (1.77-2.44),<br>p < .001 | 2.44 (2.04-2.93), p < .001 | 1.26 (1.13-1.4),<br>p < .001 | 1.67 (1.47-1.91),<br>p < .001 | Reference | 0.88 (0.78-0.98), p = .021 |
| GAD-7 | 0.97 (0.86-1.1),<br>p = .635 | 0.95 (0.83-1.09), p = .462 | 1.01 (0.93-1.1),<br>p = .797 | 0.98 (0.89-1.08),<br>p = .679 | Reference | 1.03 (0.95-1.12), p = .472 |

## DISCUSSION

This study investigated short-term trajectories of depression and anxiety symptom severity in a cohort of patients during antidepressant treatment in an outpatient behavioral health clinic. GBTM revealed six distinct depression and anxiety response trajectories. Fewer than half of patients were included in the responder groups for depressive (42.98%) and anxiety (44.89%) related symptoms over weeks 1 to 13 of antidepressant treatment. Baseline variables associated with statistically significant differences between the groups for depression included higher PHQ- 9 and C-SSRS scores related to poorer symptom response. Similarly, statistically significant differences between the anxiety groups were associated with higher GAD-7 and PSQI scores related to poorer symptom response. The highest degrees of overlap occurred between PHQ-9 and GAD-7 trajectories that follow synchronous paths (72.55%), suggesting a close relationship between symptoms of depression and anxiety. Several factors measured at baseline were also found to be uniquely associated with some trajectories (e.g., PROMIS, PSQI, SDS, and age).

Higher severity of depression and anxiety symptoms prior to antidepressant initiation was generally associated with lack of change. However, there is a small group of patients who achieved a favorable course towards remission despite initiating antidepressant therapy with anxiety and depression symptoms in the severe ranges. Other groups with moderate symptoms at baseline showed unremarkable responses. This illustrates that baseline severity is only one of a number of factors that is correlated with treatment response. Other variables associated with favorable trajectories include lower sleep health (PSQI score), younger age, lower physical and social wellbeing (PROMIS score), and lower alcohol consumption habits (AUDIT). Preliminary analysis of several individuals (n=94, 16.29%) who received psychotherapy at the site indicated similar rates of nonresponse relative to the overall sample, but rates of remission vs. response trended higher among these individuals.

Among the responder groups, there were differences in the rate of response, with at least one group demonstrating a slow initial response followed by rapid and sustained improvement in symptoms beginning in the fourth week. This may have clinical relevance in terms of determining at what point a treatment regimen should be reconsidered for a non-responding patient. This finding is juxtaposed with previous studies that suggested clinicians should change or cease treatment if no remarkable improvement was noted by the third therapy session [25, 26]. Baseline demographic and clinical variables related to the identified trajectories of symptom change can be used to predict the most likely trajectory group for treatment response, which has a potential to enhance clinical decision making, optimize resource allocation, and improve treatment outcomes [7]. Patients who are identified as potential responders to treatment may be candidates for continuing treatment, while those identified as potential nonresponders may pursue alternative interventions, such as psychotherapy, at an earlier time point [28].

Another interesting finding emerged from our analysis of patients with overlapping trajectories for anxiety and depression (Figure 3). Despite the high correlation and increased functional impairment in patients with comorbid anxiety and depression symptoms, little was known about the course of each condition when presenting together in the context of antidepressant treatment. The heatmap analysis showed that most patients had synchronous depression and anxiety trajectories after initiating treatment. In particular, patients who experienced large degrees of depressive symptomatic change similarly experienced large degrees of anxiety symptomatic change. However, there were several asynchronous cases (27.45% patients) where patients who experienced large degrees of depressive symptomatic change experienced small degrees of anxiety symptomatic change, and vice versa. One reason may be attributed to the different dosages of SSRIs sometimes necessary to achieve therapeutic benefits for anxiety as opposed to depression. However, further exploration of the causes for asynchronous change may provide some clarity regarding the interplay of depression and anxiety for patients with comorbid symptoms. A number of baseline variables were associated with symptom change or lack thereof. Older age was associated with nonresponse, which supports previous work that showed treatment resistance in older populations [27]. Poor physical, mental, and social health at baseline (as measured by PROMIS) were also associated with poorer response during antidepressant treatment. These associations were obtained based on multivariable models suggesting shared variance between these factors. Clinicians may consider these factors in their treatment plan, and more closely follow patients with these characteristics.

The findings of this study may have implications for clinical practice in antidepressant therapy and personalized clinical management of patients suffering from comorbid depression and anxiety. First, our analysis of the associations of pretreatment characteristics with symptom change may aid clinicians in making treatment recommendations. For example, the ability to identify likely nonresponders at the point of a baseline assessment might encourage clinicians and patients to pursue other treatment options earlier in the course of treatment, thereby reducing unnecessary trial-and-error, delays and risk of adverse effects. The findings might also suggest what risk factors clinicians should be more aware of in terms of evaluating treatment response trajectories. The baseline characteristics information can also be used to help anticipate likely symptom change trajectories, which can support clinical decision-making and lead to better treatment outcomes. The trajectories we described may also aid in the identification of patients whose symptoms are not improving, as well as deciding at what point clinicians should intervene or switch antidepressants or consider adjuvant treatment. On a broader scale, understanding these sub-populations can facilitate clinicians in personalizing care and management for these patients. Furthermore, our analysis showed that, although there are many features and symptoms that overlap between depression and anxiety, there are some that are unique to each. Our analysis of the several characteristics associated with the anxiety and depression trajectories derived in this study lays the groundwork for deeper investigation of the interaction and impacts that one disorder has on the other.

The results of this study should be understood within the context of some methodological limitations. First, because the analysis was limited to patients receiving antidepressant medications, it is not possible to attribute the symptom trajectories to antidepressant treatment as distinct from regression to the mean. Second, this study used data from a single outpatient mental health clinic at an urban academic medical center, which limits our ability to generalize the model’s utility across multiple different settings and health systems. It will be necessary to replicate the observed response patterns in other cohorts. Third, although commonly associated with the use of EHR data across a single health system, patients may have sought concurrent treatment at sites not affiliated with the health system and therefore not captured within our EHR. This limits our ability to fully control for or reliably measure the influence of such factors on treatment response.

## CONCLUSION

Six different trajectories of symptom change for depression and anxiety were derived. The observed trajectories included various paths to symptom response and remission that clinicians may encounter in practice, and casted two often conflated mental health conditions in their own lights. While our analysis also examined several factors associated with certain trajectory responses, deeper understanding of the underlying factors of trajectory-based subgroups may reveal insights into the mixed pathophysiology of depression and anxiety, and motivate strategies to personalize therapy for these conditions.

## Data Availability

All data produced in the present study are available upon reasonable request to the authors.

## Acknowledgements

This research is funded in part by grants from the US National Institutes of Health (grant numbers R01 GM105688, R01 MH119177, R01 MH121907, and R01 MH121922).

## Supplementary information

### 1. Characteristics of the overall sample

Across the entire sample, 42.98% of patients were characterized as responders for depressive symptoms, while 57.01% were non-responders. Baseline characteristics that significantly differed between groups included PHQ-9 and C-SSRS scores. Similar patterns emerged for anxiety symptoms, with 44.89% of patients in the responder groups, and 55.11% non- responders. Significant differences between groups were identified in the GAD-7 and PSQI scores. The types of antidepressants (SSRIs, SNRIs, TCAs, MAOIs, and atypical antidepressants) did not significantly differ across groups.

### 2. Baseline characteristics

The baseline was defined as the first session and encounter where care was established at the outpatient behavioral health clinic. Demographic variables extracted from this session included age, gender, and self-reported race. Comorbidities included post-traumatic stress disorder (PTSD), bipolar disorder, and “suicidal behavior,”, which comprised ICD-coded suicide attempt, suicidal ideation, and self-harm. The individual’s PHQ-9 and GAD-7 scores at baseline were also extracted. Several other prognostic factors were measured at baseline using multiple validated scoring instruments and questionnaires. These included the Alcohol Use Disorders Identification Test (AUDIT) which measures alcohol use and drinking behaviors; the Columbia-Suicide Severity Rating Scale (C-SSRS) measuring suicide risk; the Pittsburgh Sleep Quality Index (PSQI) which assesses sleep patterns and quality; the Patient-Reported Outcomes Measurement Information System (PROMIS) which evaluates overall physical, mental, and social wellbeing; and the Sheehan Disability Scale (SDS) which measures functional impairment in work/school, social, and family life. Lower symptom severity is indicated by lower scores on the AUDIT, C-SSRS, PSQI, and SDS questionnaires and a higher score on the PROMIS. The associations of these baseline characteristics with trajectories of depression and anxiety were investigated. All instruments were administered to the clinic patients as part of their routine clinical care.

### 3. Statistical Analysis

GBTM was employed to stratify members of the cohort into clinically meaningful subgroups with statistically similar trajectories of their depression and anxiety symptoms. GBTM has been applied in clinical contexts to reveal patient groups with similar characteristics.

Analysis was conducted in R (version 4.0.3) and GBTM was performed using the package lcmm. The number of groups and the degree of the polynomial in each of the trajectory groups were determined by using the Akaike Information Criterion (AIC) and Bayesian Information Criterion (BIC) to measure model fit with lower AIC and BIC values indicating better fit.

After identifying trajectory groups using GBTM, further univariate and multivariate tests were performed to obtain associations between baseline prognostic factors and trajectory groups. In particular, the continuous characteristics (PHQ-9, GAD-7, AUDIT, CSSRS,

PROMIS, PSQI, and SDS) were described using the mean and standard deviation (SD), while categorical characteristics (e.g., race and gender) were described using frequency counts. For the univariate analyses, we applied a student’s t-test, Mann-Whitney U test, Chi- square test, and Fisher’s exact test where appropriate. Analysis of covariance (ANCOVA) for the between-trajectory group comparisons was also employed on the basis of the general linear model (GLM), adjusting for age at baseline. For the multivariate analyses, multinomial logistic regression models were used to assess associations of baseline prognostic factors with trajectory group membership, including their ability to predict membership based on pre-treatment patient characteristics.

### 4. Characteristics of the derived trajectory groups on anxiety symptom change

There were also six trajectory groups reflecting pathways of anxiety symptom change, which included high, nonresponse (Cluster 1), high, response (Cluster 2), moderate, nonresponse-1 (Cluster 3), moderate, nonresponse-2 (Cluster 4), high, remission (Cluster 5), and moderate, remission (Cluster 6). Patients in the trajectory group high, nonresponse underwent limited change during antidepressant treatment. Patients in the high, response group demonstrated a substantial decrease in anxiety symptoms. Patients in the high, remission group had severe anxiety at baseline but demonstrated the greatest symptom improvement and ultimately achieved levels below typical remission criteria (GAD-7 ≤ 5). Patients in the moderate, remission group initiated care with moderate anxiety symptoms and displayed steady improvement, also reaching levels below typical remission criteria. Although patients in high, remission and moderate, remission groups achieved the largest symptom improvements, the slope of high, remission group was steeper than that of moderate, remission group, which suggests that the speed of improvement was higher in high, remission group. We observed unremarkable improvement for patients in GAD-7 trajectory groups moderate, nonresponse-1 and moderate, nonresponse-2 groups. Patient characteristics for the GAD-7 trajectory groups are shown in eTable 1.

### 5. The associations of baseline patient characteristics with GAD-7 trajectory group membership

In terms of the GAD-7 trajectories, relative to being in GAD-7 trajectory Cluster 5 (High, remission), the probability of being in Cluster 1 (High, nonresponse) was higher and was associated with poorer overall health (lower PROMIS scores); the probability of being in GAD-7 trajectory Cluster 2 (High, response) was higher and was associated with more severity of anxiety prior to treatment (higher baseline GAD-7 scores). The probability of being in GAD-7 trajectory Cluster 3 (Moderate, nonresponse1) was higher and was associated with less severity of anxiety prior to treatment (lower baseline GAD-7 scores); the probability was also higher if patients’ race was non-white. The probability of being in GAD- 7 trajectory Cluster 4 (Moderate, nonresponse2) was higher and was associated with less severity of anxiety prior to treatment and poorer sleep quality (lower baseline GAD-7 scores and higher PSQI); the probability was also higher if patients’ race was non-white. Finally, the probability of being in GAD-7 trajectory Cluster 6 (Moderate, remission) was higher and was associated with older, less severity of anxiety prior to treatment, and better overall health (higher age, PROMIS, and lower baseline GAD-7 scores).

**eFigure 1:**
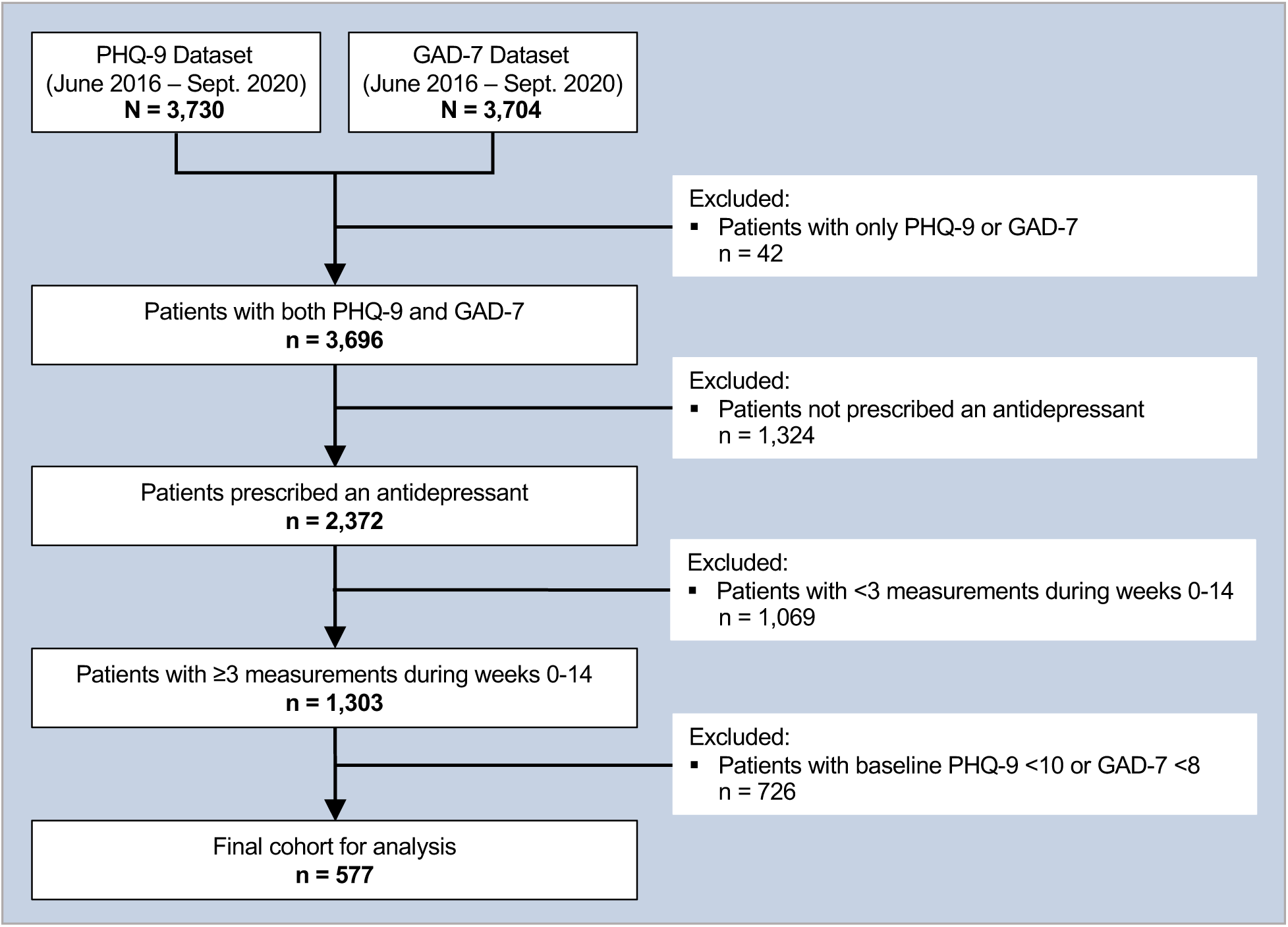
The inclusion/exclusion cascade for identifying study participants.

**eTable 1:** Patient characteristics for the GAD-7 trajectory groups.

**7. eTable 1: Patient characteristics for the GAD-7 trajectory groups**
| Characteristic | All patients | Cluster 1<br>(High,<br>nonresponse) | Cluster 2<br>(High,<br>response) | Cluster 3<br>(Moderate,<br>nonresponse1) | Cluster 4<br>(Moderate,<br>nonresponse2) | Cluster 5<br>(High,<br>remission) | Cluster 6<br>(Moderate,<br>remission) | *P* value |
| --- | --- | --- | --- | --- | --- | --- | --- | --- |
| Age,<br>mean (SD) | 39.34<br>(12.87) | 42.02 (12.9) | 37.05<br>(12.36) | 40.35 (13.5) | 37.65 (13.75) | 37.41<br>(10.67) | 41.14<br>(12.03) | 0.006595144 |
| PHQ-9,<br>mean (SD) | 16.19<br>(4.26) | 18.43 (4.34) | 16.98<br>(4.31) | 15.52 (4.08) | 14.81 (3.93) | 15.95<br>(3.92) | 15.35<br>(3.72) | 2.72E-09 |
| GAD-7,<br>mean (SD) | 15.35<br>(3.81) | 18.51 (2.19) | 17.96<br>(2.01) | 14.92 (2.8) | 12.14 (2.63) | 17.64<br>(2.45) | 11.72<br>(3.29) | 3.87E-65 |
| AUDIT,<br>mean (SD) | 3.69<br>(1.61) | 3.71 (1.65) | 3.58<br>(1.76) | 3.94 (1.58) | 3.46 (1.49) | 3.94<br>(1.72) | 3.65 (1.38) | 0.371474188 |
| C-SSRS,<br>mean (SD) | 1.47<br>(1.2) | 1.69 (1.32) | 1.5<br>(1.16) | 1.46 (1.19) | 1.34 (1.19) | 1.45<br>(1.02) | 1.4 (1.2) | 0.253010892 |
| PROMIS,<br>mean (SD) | 13.57<br>(1.89) | 12.73 (1.85) | 13.54<br>(1.93) | 13.66 (1.73) | 13.84 (1.68) | 13.59<br>(2.23) | 14.1 (1.69) | 9.27E-07 |
| PSQI,<br>mean (SD) | 10.9<br>(2.7) | 11.92 (2.61) | 10.52<br>(2.88) | 11.14 (2.44) | 10.57 (2.89) | 10.8<br>(2.28) | 10.37<br>(2.58) | 1.11E-05 |
| SDS,<br>mean (SD) | 19.56<br>(4.72) | 21.48 (5.11) | 20.35<br>(4.86) | 19.13 (4.64) | 18.29 (3.94) | 20.36<br>(4.17) | 18.0 (4.34) | 8.43E-15 |
| Gender – Female<br>(%) | 373<br>(64.64) | 70 (66.04) | 63<br>(63.00) | 64 (65.98) | 72 (62.61) | 42 (63.64) | 62 (66.67) | 0.983607073 |
| Race – White<br>(%) | 313<br>(54.25) | 58 (54.72) | 52<br>(52.00) | 50 (51.55) | 57 (49.57) | 42 (63.64) | 54 (58.06) | 0.492842977 |

**eTable 2:** Associations of baseline patient characteristics with PHQ-9 trajectory group membership.

8. eTable 2: Associations of baseline patient characteristics with PHQ-9 trajectory group membership
| Characteristic | GAD-7 Class 1<br>(High,<br>nonresponse):<br>RR (95%CI) &<br>p-value | GAD-7 Class 2<br>(High,<br>response):<br>RR (95%CI) &<br>p-value | GAD-7 Class 3<br>(Moderate,<br>nonresponse1):<br>RR (95%CI) &<br>p-value | GAD-7 Class 4<br>(Moderate,<br>nonresponse2):<br>RR (95%CI) & p-value | GAD-7 Class 5<br>(High,<br>remission):<br>RR (95%CI)<br>& p-value | GAD-7 Class 6<br>(Moderate,<br>remission):<br>RR (95%CI) &<br>p-value |
| --- | --- | --- | --- | --- | --- | --- |
| Age | 1.01 (0.98-1.04), p = .402 | 1.0 (0.97-1.02), p = .763 | 1.02 (1.0-1.05), p = .106 | 1.01 (0.98-1.04), p = .361 | Reference | 1.04 (1.01-1.07), p = .012 |
| Gender – Female | 0.88 (0.45-1.72), p = .715 | 0.93 (0.48-1.81), p = .838 | 1.17 (0.58-2.36), p = .661 | 0.96 (0.46-2.01), p = .911 | Reference | 1.1 (0.5-2.42), p = .806 |
| Race – Non-White | 1.28 (0.66-2.47), p = .47 | 1.6 (0.83-3.07), p = .16 | 2.05 (1.03-4.08), p = .04 | 2.47 (1.18-5.16), p = .016 | Reference | 1.99 (0.92-4.32), p = .082 |
| AUDIT | 0.95 (0.79-1.15), p = .621 | 0.85 (0.69-1.04), p = .105 | 1.06 (0.87-1.29), p = .588 | 0.88 (0.7-1.1), p = .261 | Reference | 0.91 (0.72-1.16), p = .463 |
| C-SSRS | 0.84 (0.61-1.16), p = .282 | 0.88 (0.63-1.24), p = .471 | 0.91 (0.65-1.28), p = .603 | 0.76 (0.52-1.11), p = .162 | Reference | 0.79 (0.52-1.19), p = .256 |
| LEC_5 | 1.03 (0.95-1.13), p = .47 | 1.06 (0.98-1.16), p = .163 | 0.99 (0.9-1.08), p = .749 | 1.0 (0.91-1.1), p = .977 | Reference | 0.97 (0.87-1.08), p = .597 |
| PCL-C | 1.1 (1.0-1.21), p = .053 | 0.99 (0.9-1.08), p = .784 | 1.07 (0.97-1.18), p = .196 | 1.1 (0.99-1.22), p = .09 | Reference | 1.03 (0.91-1.15), p = .664 |
| PROMIS | 0.82 (0.72-0.94), p < .001 | 1.01 (0.89-1.15), p = .895 | 1.19 (1.05-1.36), p < .001 | 1.5 (1.31-1.73), p < .001 | Reference | 1.53 (1.33-1.77), p < .001 |
| PSQI | 1.0 (0.87-1.16), p = .964 | 0.88 (0.77-1.02), p = .081 | 1.2 (1.04-1.39), p = .016 | 1.21 (1.03-1.42), p = .019 | Reference | 1.15 (0.97-1.37), p = .1 |
| SDS | 0.94 (0.87-1.02), p = .121 | 0.98 (0.91-1.05), p = .517 | 0.98 (0.91-1.07), p = .7 | 1.02 (0.94-1.12), p = .6 | Reference | 1.02 (0.93-1.12), p = .702 |
| PHQ-9 | 1.1 (1.0-1.22), p = .054 | 1.1 (1.0-1.22), p = .05 | 1.06 (0.95-1.17), p = .302 | 1.13 (1.01-1.26), p = .036 | Reference | 1.19 (1.05-1.34), p < .001 |
| GAD-7 | 1.03 (0.89-1.2), p = .692 | 1.06 (0.92-1.23), p = .424 | 0.65 (0.56-0.76), p < .001 | 0.48 (0.41-0.57), p < .001 | Reference | 0.46 (0.39-0.55), p < .001 |

**eTable 3:** The process for choosing the best number of groups based on PHQ-9 score in GBTM with AIC and BIC.

9. eTable 3: The process for choosing the best number of groups based on PHQ-9 score in GBTM with AIC and BIC
| Number of Groups | Akaike Information Criterion (AIC) | Bayesian Information Criterion (BIC) | Class distribution (# and %) |
| --- | --- | --- | --- |
| 2 | 48077.37 | 48125.46 | class1 class2<br>N 206 379<br>% 35.21 64.79 |
| 3 | 47098.54 | 47159.74 | class1 class2 class3<br>N 151 264 170<br>% 25.81 45.13 29.06 |
| 4 | 46660.38 | 46734.69 | class1 class2 class3 class4<br>N 56 147 254 128<br>% 9.57 25.13 43.42 21.88 |
| 5 | 46424.13 | 46511.56 | class1 class2 class3 class4 class5<br>N 59 103 150 114 159<br>% 10.09 17.61 25.64 19.49 27.18 |
| 6 | **46278.78** | **46379.33** | **class1 class2 class3 class4 class5 class6**<br>**N 45 111 109 133 113 74**<br>**% 7.69 18.97 18.63 22.74 19.32 12.65** |
| 7 | 46280.94 | 46394.6 | class1 class2 class3 class4 class5 class6 class7<br>N 36 95 145 139 75 95 0<br>% 6.15 16.24 24.79 23.76 12.82 16.24 0 |

**eTable 4:** The process for choosing the best number of groups based on GAD-7 score in GBTM with AIC and BIC.

**10. eTable 4: The process for choosing the best number of groups based on GAD-7 score in GBTM with AIC and BIC**
| Number of Groups | Akaike Information Criterion (AIC) | Bayesian Information Criterion (BIC) | Class distribution (# and %) |
| --- | --- | --- | --- |
| 2 | 47313.76 | 47339.99 | class1 class2<br>N 276 309<br>% 47.18 52.82 |
| 3 | 46515.1 | 46554.45 | class1 class2 class3<br>N 132 248 205<br>% 22.56 42.39 35.04 |
| 4 | 46227.12 | 46279.58 | class1 class2 class3 class4<br>N 87 193 201 104<br>% 14.87 32.99 34.36 17.78 |
| 5 | 46006.89 | 46072.47 | class1 class2 class3 class4 class5<br>N 90 127 161 114 93<br>% 15.38 21.71 27.52 19.49 15.9 |
| **6** | **45908.99** | **45987.68** | **class1 class2 class3 class4 class5 class6**<br>**N 87 118 97 106 100 77**<br>**% 14.87 20.17 16.58 18.12 17.09 13.16** |
| 7 | 45914.99 | 46006.8 | class1 class2 class3 class4 class5 class6 class7<br>N 87 118 97 106 100 77 0<br>% 14.87 20.17 16.58 18.12 17.09 13.16 0 |

## REFERENCES

1. Gaspersz, R., et al., Patients with anxious depression: overview of prevalence, pathophysiology and impact on course and treatment outcome. Current opinion in psychiatry, 2018. 31(1): p. 17–25.

2. Hung, C.-I., et al., Comorbidity with more anxiety disorders associated with a poorer prognosis persisting at the 10-year follow-up among patients with major depressive disorder. Journal of Affective Disorders, 2020. 260: p. 97–104.

3. Jha, M.K. and M.H. Trivedi, Personalized antidepressant selection and pathway to novel treatments: clinical utility of targeting inflammation. International journal of molecular sciences, 2018. 19(1): p. 233.

4. Leuchter, A.F., et al., A new paradigm for the prediction of antidepressant treatment response. Dialogues in clinical neuroscience, 2009. 11(4): p. 435.

5. Queen, A.H., D.H. Barlow, and J. Ehrenreich-May, The trajectories of adolescent anxiety and depressive symptoms over the course of a transdiagnostic treatment. Journal of anxiety disorders, 2014. 28(6): p. 511–521.

6. Rossom, R.C., et al., Predictors of poor response to depression treatment in primary care. Psychiatric Services, 2016. 67(12): p. 1362–1367.

7. Saunders, R., et al., Trajectories of depression and anxiety symptom change during psychological therapy. Journal of affective disorders, 2019. 249: p. 327–335.

8. Smagula, S.F., et al., Antidepressant response trajectories and associated clinical prognostic factors among older adults. JAMA psychiatry, 2015. 72(10): p. 1021–1028.

9. Sunderland, M., et al., Investigating trajectories of change in psychological distress amongst patients with depression and generalised anxiety disorder treated with internet cognitive behavioural therapy. Behaviour research and therapy, 2012. 50(6): p. 374–380.

10. Uher, R., et al., Trajectories of change in depression severity during treatment with antidepressants. Psychological medicine, 2010. 40(8): p. 1367–1377.

11. Frank, E., et al., Conceptualization and rationale for consensus definitions of terms in major depressive disorder: remission, recovery, relapse, and recurrence. Archives of general psychiatry, 1991. 48(9): p. 851–855.

12. Mori, M., H.M. Krumholz, and H.G. Allore, Using latent class analysis to identify hidden clinical phenotypes. Jama, 2020. 324(7): p. 700–701.

13. Hunter, A.M., et al., Antidepressant response trajectories and quantitative electroencephalography (QEEG) biomarkers in major depressive disorder. Journal of psychiatric research, 2010. 44(2): p. 90–98.

14. Cui, X., et al., Outcomes and predictors of late-life depression trajectories in older primary care patients. The American Journal of Geriatric Psychiatry, 2008. 16(5): p. 406–415.

15. Kroenke, K., et al., Anxiety disorders in primary care: prevalence, impairment, comorbidity, and detection. Annals of internal medicine, 2007. 146(5): p. 317–325.

16. Saunders, J.B., et al., Development of the alcohol use disorders identification test (AUDIT): WHO collaborative project on early detection of persons with harmful alcohol consumption-II. Addiction, 1993. 88(6): p. 791–804.

17. Posner, K., et al., Columbia-suicide severity rating scale (C-SSRS). New York, NY: Columbia University Medical Center, 2008. 10.

18. Buysse, D.J., et al., The Pittsburgh Sleep Quality Index: a new instrument for psychiatric practice and research. Psychiatry research, 1989. 28(2): p. 193–213.

19. Hays, R.D., et al., Development of physical and mental health summary scores from the patient-reported outcomes measurement information system (PROMIS) global items. Quality of Life Research, 2009. 18(7): p. 873–880.

20. Leon, A.C., et al., Assessing psychiatric impairment in primary care with the Sheehan Disability Scale. Int J Psychiatry Med, 1997. 27(2): p. 93–105.

21. Wu, S., et al., Association of trajectory of cardiovascular health score and incident cardiovascular disease. JAMA network open, 2019. 2(5): p. e194758–e194758.

22. Proust-Lima, C., et al., Package ‘lcmm’. 2022.

23. Sakamoto, Y., M. Ishiguro, and G. Kitagawa, Akaike information criterion statistics. Dordrecht, The Netherlands: D. Reidel, 1986. 81(10.5555): p. 26853.

24. Vrieze, S.I., Model selection and psychological theory: a discussion of the differences between the Akaike information criterion (AIC) and the Bayesian information criterion (BIC). Psychological methods, 2012. 17(2): p. 228.

25. Lambert, M.J., Outcome in psychotherapy: the past and important advances. 2013.

26. Romera, I., et al., Early switch strategy in patients with major depressive disorder: a double-blind, randomized study. Journal of clinical psychopharmacology, 2012. 32(4): p. 479–486.

27. Reynolds III, C.F. and D.J. Kupfer, Depression and aging: a look to the future. Psychiatric Services, 1999. 50(9): p. 1167–1172.

28. Al-Harbi, Khalid Saad. Treatment-resistant depression: therapeutic trends, challenges, and future directions. Patient preference and adherence, 2012. 6: p. 369–388.

